# AI learns racial information from the values of vital signs

**DOI:** 10.1101/2023.12.11.23299819

**Authors:** Bojana Velichkovska, Hristijan Gjoreski, Daniel Denkovski, Marija Kalendar, Irene Dankwa Mullan, Judy Wawira Gichoya, Nicole Martinez, Leo Anthony Celi, Venet Osmani

**Affiliations:** Faculty of Electrical Engineering and Information Technologies, Cyril and Methodius University, Skopje, North Macedonia; Milken Institute School of Public Health, George Washington University, Washington, DC, USA; Department of Radiology and Imaging Sciences, Emory University School of Medicine, Atlanta, GA, USA; Center for Biomedical Ethics, Stanford University School of Medicine, Stanford, CA, USA; Division of Pulmonary, Critical Care and Sleep Medicine, Beth Israel Deaconess Medical Center, Boston, MA, USA; Institute of Medical Engineering and Science, Massachusetts Institute of Technology, Boston, MA, USA; Department of Biostatistics, Harvard T.H. Chan School of Public Health, Boston, Massachusetts, USA; Information School, University of Sheffield, UK

## Abstract

**Background:** Bias in medical practice is multifaceted, including treatment variations across race-ethnicity, unconscious bias in healthcare providers’ attitudes, and bias in clinical scores. However, far less is known about the potential racial bias in routinely collected, essential information in clinical decision-making, namely vital signs.

**Research question:** Do vital signs embed racial information that can be learned by AI algorithms?

**Study Design and Methods:** Retrospective cohort study of critically ill patients between 2014 and 2015 from the multi-centre eICU-CRD critical care database involving 335 Intensive Care Units (ICU) based in 208 US hospitals, containing 200,859 patient admissions. We extracted 10,763 critical care admissions of patients aged 18 and over, alive during the first 24 hours after admission to ICU with recorded self-reported race as well as at least two measurement of heart rate, oxygen saturation, respiratory rate, and blood pressure. Pairs of racial subgroups were matched based on age, gender, admission diagnosis and APACHE IV scores. Traditional machine learning algorithms, including XGBoost and Logistic regression were used to predict self-reported race using values of vital signs as an input.

**Results:** AI models derived from only six vital signs can predict patients’ self-reported race with an AUC of 0.74 (± 0.022) between White and Black patients. Technologies used to measure oxygen saturation are a significant source of self-reported racial information (AUC of 0.72 ± 0.028), in addition to blood pressure measurements (AUC of 0.63 ± 0.035). Care delivery practices do not present a significant source of racial information (AUC of 0.57 ± 0.019). However, even when controlling for these known factors, self-reported race can still be learned from vital signs, whose origin we cannot currently explain.

**Interpretation:** Vital signs embed racial information that can be learned by AI algorithms, posing a significant risk to equitable clinical decision-making. Mitigating measures might be challenging, considering fundamental role of vital signs.

## 1 Introduction

As a subset of AI, machine learning (ML) algorithms are evolving to tackle increasingly complex clinical challenges ^1–3^. A general appeal is that clinical practice is likely to benefit from algorithmic assisted decision-making by optimising clinical workflows, diagnostic interventions, and enhancing personalised precision care. It is likely that some of the insights derived from these algorithms will assist in clinical decision-making where patients’ lives are at risk. Therefore, it is essential that ML algorithms that become integrated as part of decision-making in clinical practice be robust, reliable, and unbiased.

Health care disparities resulting from discrimination and bias are pervasive. These can be encoded in algorithms trained on clinical data captured in electronic health records. Existing inequities can be perpetuated or even magnified by algorithms developed to inform decision-making because of bias in the data used to develop the models, bias introduced during the model development, or bias during deployment and post-deployment monitoring. Any of these can result in decision-making that can be discriminatory and harmful to socially disadvantaged population groups that are not adequately represented in the data.

Treatment variation across race-ethnicity that is not explained by patient or disease factors has been detailed in several studies, accompanied by significant evidence of unconscious bias in healthcare providers’ attitudes, expectations, and behaviour ^4–6^. The presence of this type of bias in medical practice is further amplified if the discriminatory attitudes and behaviours are in turn modelled as disease mechanisms or as decision support algorithms that are implemented by care providers. This phenomenon is described by Brooks in an opinion piece that framed unconscious bias as a “silent curriculum” ^7^. Furthermore, a recent study illustrates racial bias in the descriptions of patients’ electronic health records (EHR), showing that Black patients are 2.5 times more likely to have one or more negative descriptors in their EHR compared with White patients ^8^.

Bias embedded in data has been illustrated by Obermeyer et al. where “at a given risk score, Black patients are considerably sicker than White patients, as evidenced by signs of uncontrolled illnesses.”. The algorithm learned to predict care costs, placing Black patients in the same risk category as a subset of White patients, while having considerably worse symptoms ^9^. To add to the severity of the problem, the number of Black patients who should have been referred for complex care was reduced by half based on the recommendation of the ML algorithm.

While there is awareness of bias in clinical scores, far less is known about the potential racial bias in routinely collected, essential information in clinical decision-making, namely vital signs. Therefore, we sought to investigate whether self-reported race can be learned from routine measurements of vital signs. If sensitive attributes such as race-ethnicity are easily learned from essential clinical data, then it is of significant concern if these attributes become an embedded part of clinical decision-making and treatment optimisation.

## 2 Methods

This study followed the Strengthening the Reporting of Observational Studies in Epidemiology (STROBE) reporting guidelines ^10^. Given that our patient cohort is made up of predominantly White patients, we devised a matching cohort to mitigate representation bias of other races. We matched patients of different races based on admission diagnosis, gender, age, and Acute Physiology and Chronic Health Evaluation (APACHE IV) score to equally represent the three population subgroups considered in our study, namely Black, Hispanic, and White patients. Classical ML algorithms (Logistic Regression (LR) and XGBoost ^11^) were then used to investigate whether the patients’ self-reported race can be predicted using solely patients’ vital signs.

### 2.1 Clinical data sources and study population

We used the eICU Collaborative Research Database (eICU-CRD) containing 200,859 admissions collected from 335 ICUs across 208 hospitals in the United States (US) admitted between 2014 and 2015 ^12^. We selected all adult patients (age 18 and over) that were alive within the first 24 hours after ICU admission and had at least one clinically valid measurement for all the vital signs considered for this study: heart rate, SpO2, SaO2, respiratory rate, non-invasive, and invasive blood pressure (systolic, diastolic, and mean). There were insufficient measurements of temperature. We selected data from the first 24 hours, given that a patient’s presenting vital signs inform follow-up and life-saving interventions. This means that potentially biased data during this critical period have the potential to carry most harm. We extracted mean, minimum, maximum, and variance of the selected vital signs. Patients with missing APACHE IV score, admission diagnosis, age, or gender were excluded from the study. Inclusion of the three dominant racial groups resulted in a total of 10,763 patients, of which 9,215 (85%) were White, 1,066 (10%) were Black, and 482 (5%) were Hispanic. Since significantly less data was available for Asian American and Native American patients (177 and 84 respectively) they were not included in this study.

Cohort matching resulted in 844 Black patients matched to the same number of White patients, 392 Hispanic patients matched to the same number of White patients, and 222 Hispanic patients matched to the same number of Black patients.

### 2.2 Statistical analysis

Baseline characteristics of the patients were analysed using medians (IQRs) for continuous variables and frequencies (percentages) for categorical variables. We used the Kruskal–Wallis test (one-way ANOVA) for continuous variables and the chi-square test for categorical variables to compare different racial and ethnic subgroups. Due to the selection criteria (shown in Appendix 2) no patients with missing data remained.

### 2.3 Model development and validation

We analysed binary outcomes, namely whether vital signs can predict self-reported races of Black versus White. We carried out the same analysis for Hispanic and White patients, and Hispanic and Black patients. We used two ML algorithms to derive the models and evaluate their performance, namely Logistic Regression (LR) and XGBoost^11^. As XGBoost is prone to overfitting, we evaluated two versions of the XGBoost algorithm: version with the default parameters, and an optimised version (with parameters selected using random search^13^). We also evaluated a shallow neural network, which did not result in a significant performance improvement.

We used stratified 5-fold cross-validation to evaluate the models, meaning the data were divided into 5 folds so each fold maintains the original distribution class-wise. Model derivation was performed on 4 folds, while the remaining fold was used to validate the model’s performance. We repeated this process 5 times, for each of the folds, and the final results were averaged over all folds. We assessed the performance of our models by computing the area under the receiver operator characteristic curve (AUC-ROC).

## 3 Results

Out of 10,763 patients, 1,688 met the inclusion criteria for the first matched cohort (Black and White patients), 784 for the second pair of matched cohorts (Hispanic and White patients) and 444 for the third matched cohort (Hispanic and Black patients). The patient baseline characteristics for each of the matched cohorts are summarised in Table 1. Initially, we investigated prediction of self-reported race using heart rate, SpO2, SaO2, respiratory rate, and blood pressure (systolic, diastolic, and MAP measured through arm cuff as well as invasively). Then, we performed sensitivity analysis, focusing solely on patients with known comorbidities.

**Table 1.** Patient characteristics for each of the matched cohorts from the eICU-CRD database.

|  | Black and White matched patient cohort |  |  | Hispanic and White matched patient cohort |  |  | Hispanic and Black matched patient cohort |  |  |
| --- | --- | --- | --- | --- | --- | --- | --- | --- | --- |
| Clinical values | Black | White | p-value | Hispanic | White | p-value | Hispanic | Black | p-value |
| Patients | 844 | 844 | - | 392 | 392 | - | 222 | 222 | - |
| Gender (male) | 457 (51.7%) | 483 (57.23%) | - | 236 (60.2%) | 238 (60.7%) | - | 141 (63.5%) | 127 (57.21%) | - |
| Age | 60 [51, 69] | 65 [56, 73] | < 0.05 | 63 [53, 73] | 66 [57, 73] | < 0.05 | 63 [54, 73] | 62 [52, 70] | 0.151 |
| Heart rate | 87 [75, 99] | 84 [73, 97] | < 0.05 | 84 [73, 97] | 83 [73, 96] | < 0.05 | 84 [74, 96] | 86 [76, 98] | < 0.05 |
| Invasive Oxygen Saturation | 97.6 [95, 99] | 97.4 [95, 99] | < 0.05 | 97 [95, 99] | 97 [95, 99] | 0.07 | 97 [95, 99] | 97 [95, 99] | < 0.05 |
| Oxygen Saturation (Pulse Ox) | 99 [97, 100] | 98 [95, 99] | < 0.05 | 98 [96, 100] | 97 [95, 99] | < 0.05 | 98 [96, 100] | 99 [96, 100] | < 0.05 |
| Respiration rate | 19 [15, 24] | 19 [16, 23] | < 0.05 | 19 [16, 23] | 19 [15, 23] | < 0.05 | 19 [16, 23] | 19 [15, 24] | < 0.05 |
| Invasive Systolic BP | 123 [107, 141] | 120 [106, 138] | < 0.05 | 122 [108, 138] | 123 [108, 140] | < 0.05 | 119 [106, 135] | 122 [108, 140] | < 0.05 |
| Invasive Diastolic BP | 61 [53, 70] | 58 [50, 66] | < 0.05 | 59 [51, 68] | 59 [51, 68] | < 0.05 | 58 [51, 67] | 60 [53, 68] | < 0.05 |
| Invasive Mean BP | 80 [72, 91] | 78 [69, 87] | < 0.05 | 79 [70, 90] | 79 [70, 90] | < 0.001 | 78 [69, 88] | 80 [72, 89] | < 0.05 |
| Systolic blood pressure | 121 [105, 139] | 118 [103, 135] | < 0.05 | 120 [106, 135] | 119 [105, 136] | < 0.001 | 117 [104, 133] | 123 [106, 141] | < 0.05 |
| Diastolic blood pressure | 66 [57, 76] | 63 [55, 73] | < 0.05 | 64 [55, 74] | 63 [55, 73] | < 0.05 | 62 [54, 72] | 65 [56, 75] | < 0.05 |
| Mean Arterial Pressure (MAP) | 82 [72, 94] | 79 [69, 91] | < 0.05 | 78 [69, 89] | 80 [70, 91] | < 0.05 | 76 [68, 87] | 82 [72, 94] | < 0.05 |

### 3.1 Vital signs as a source of bias

Initially, we investigated the presence of racial bias in the overall patient cohort and use this as a baseline measure. Our analysis of 1,688 patient admissions reveals that the XGBoost algorithm can predict patients’ self-reported race using only vital signs, with a performance of AUC 0.74 (± 0.022) as shown in Figure 1 and further detailed in Appendix 1.

**Figure 1.**
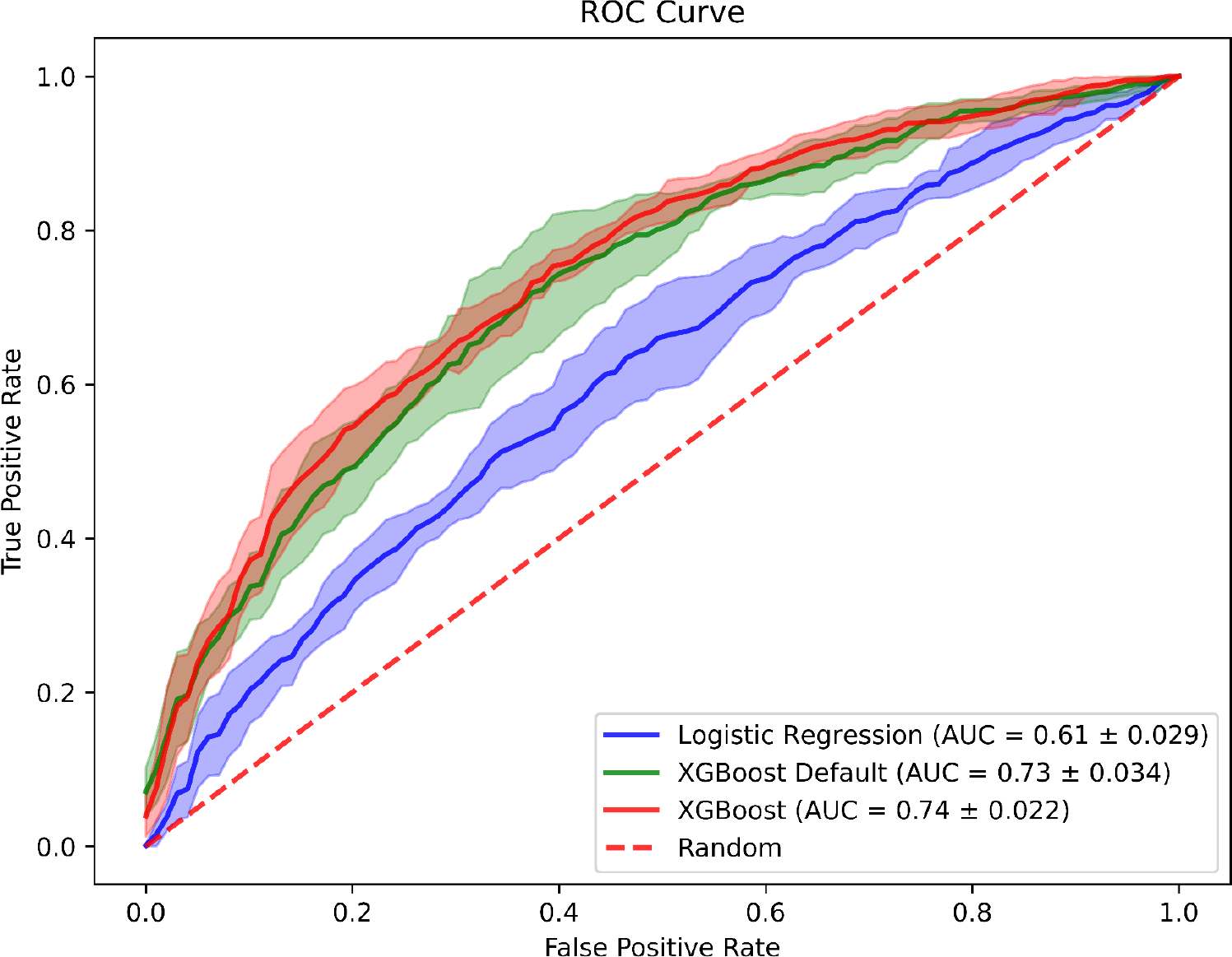
Performance of machine learning models in predicting patients’ self-reported race using vital signs only as input for Black and White patients. Analysis is based on Logistic Regression, XGBoost with default parameters and XGBoost with optimised parameters found using random search.

Furthermore, similar performance in predicting patients’ race also holds when considering patients with comorbidities, such as sepsis, essential hypertension, acute kidney failure, and chronic kidney disease, except in patients with heart failure, as shown in Table 2.

**Table 2.** Prediction of race from six vital signs in patients with comorbidities for Logistic Regression (LR), XGBoost, and XGBoost with optimised (opt) hyperparameters. Results are shown using Area Under the receiver operating characteristic Curve (AUC) along with standard deviation (SD).

|  | Sepsis | Essential hypertension | Heart failure | Acute kidney failure | Chronic kidney disease |
| --- | --- | --- | --- | --- | --- |
| LR | $0.57 \pm 0.176$ | $0.62 \pm 0.073$ | $0.53 \pm 0.113$ | $0.60 \pm 0.049$ | $0.57 \pm 0.099$ |
| XGBoost | $0.69 \pm 0.093$ | $0.60 \pm 0.064$ | $0.51 \pm 0.050$ | $0.71 \pm 0.068$ | $0.72 \pm 0.079$ |
| XGBoost (opt) | $0.71 \pm 0.077$ | $0.60 \pm 0.091$ | $0.50 \pm 0.063$ | $0.69 \pm 0.81$ | $0.67 \pm 0.064$ |

Considering these results, we then probed possible origins of racial information in vital signs through variable saliency analysis. We used SHAP method to understand the influence of each of the vital signs in predicting patients’ self-reported race. The SHAP analysis, shown in Figure 2 revealed that oxygen saturation measured through pulse oximetry, was the most influential variable in predicting patients’ race. Therefore, our analysis focused on further investigating not only pulse oximetry but also technological approaches used to measure vital signs in general as potential sources of racial information.

**Figure 2.**
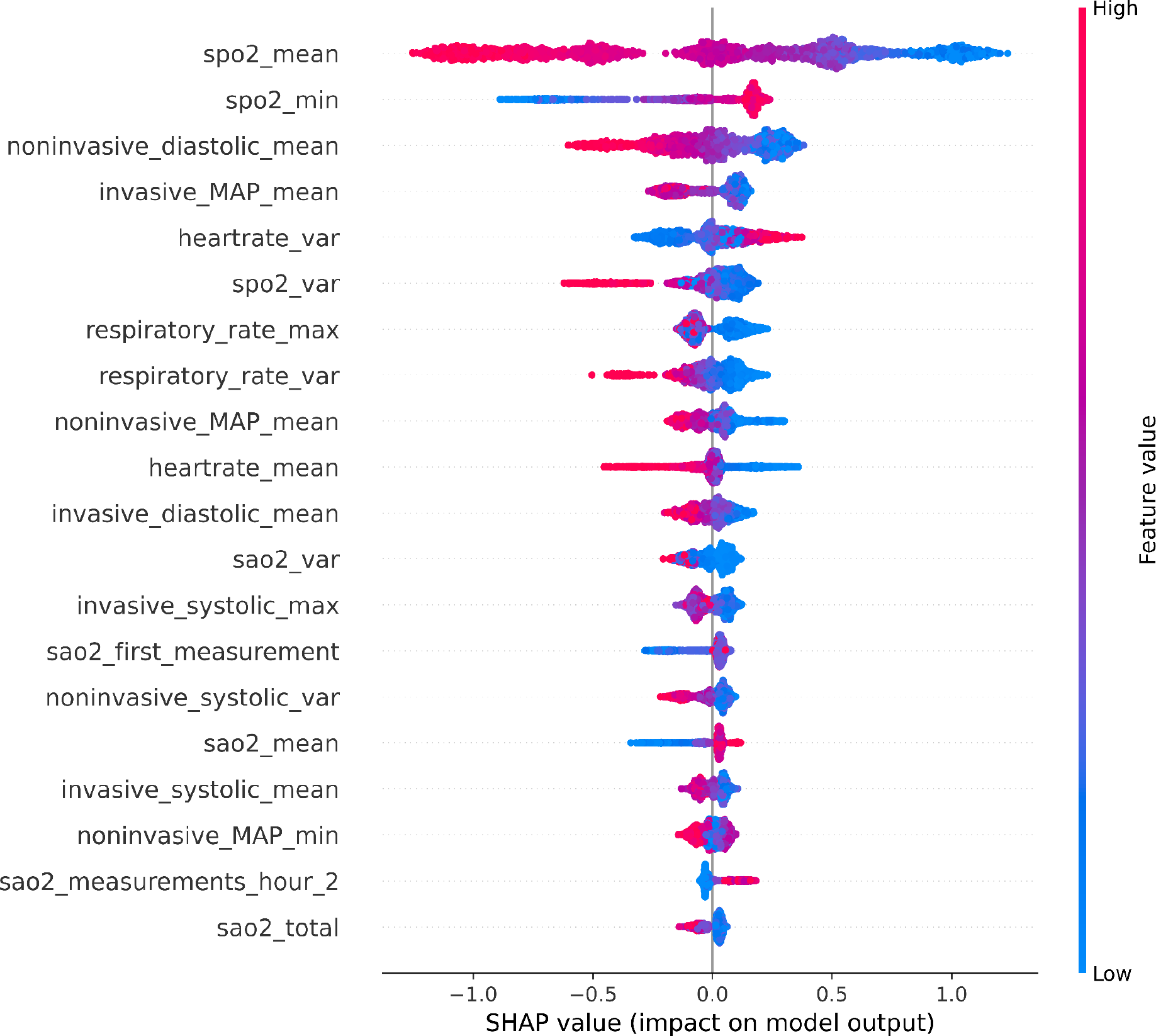
Importance of variables in predicting patients’ race using all the six vital signs in Black and White patients. Variables are shown in order of importance from the top with pulse oximetry being the most influential in predicting patients’ race. The variable name suffix indicates whether mean (_mean), minimum (_min), maximum (_max) or variance (_var) value was used for a particular vital sign variable. The frequencies of measurement were considered in the entire 24 hours (_total), during each of the first six hours, after admission (hour_[1-6]). The maximum measurements taken during a period of 1 hour are labelled as maximum_in_24_hours.

### 3.2 Technologies used to measure vital signs as a potential source of racial information

We investigated whether the devices used to measure vital signs can be a source of self-reported racial information since earlier work has shown that differences in skin colour can influence pulse oximetry readings^14^. Furthermore, acquiring accurate readings of blood pressure using an arm cuff is challenging in patients with high BMI^15^. Therefore, we investigated whether ML algorithms could pick up these known differences. We divided the results below into investigating i) potential racial information in blood pressure measurements using an arm cuff and ii) potential racial information in oxygen saturation measurements using pulse oximetry.

#### 3.2.1 Racial information in blood pressure values measured using an arm cuff

We compared performance of self-reported race prediction using blood pressure values measured through an arm cuff (non-invasive) with that of using an arterial line (invasive), in addition to using both types of measurements.

In contrast to the existing literature, our results show that while there is a presence of racial information in blood pressure measurements irrespective of the measurement method used (AUC of 0.63 ± 0.035), we did not find major differences between values of blood pressure using an arm cuff (AUC of 0.64 ± 0.036) in comparison to the values measured through an arterial line (AUC of 0.63 ± 0.025). These results are summarised in Table 3.

**Table 3.** Race prediction performance results for different potential sources of bias using Logistic Regression (LR), XGBoost, and XGBoost with optimised (opt) hyperparameters. Results are shown using Area Under the receiver operating characteristic Curve (AUC) along with standard deviation (SD).

|  | Non-invasive blood pressure | Invasive blood pressure | Non-invasive and invasive blood pressure | Non-invasive oxygen saturation | Invasive oxygen saturation | Non-invasive and invasive oxygen saturation | Frequency of measurements of vital signs | Other sources of bias |
| --- | --- | --- | --- | --- | --- | --- | --- | --- |
| LR | $0.61 \pm 0.032$ | $0.58 \pm 0.027$ | $0.59 \pm 0.024$ | $0.73 \pm 0.033$ | $0.59 \pm 0.032$ | $0.72 \pm 0.032$ | $0.55 \pm 0.043$ | $0.62 \pm 0.044$ |
| XGBoost | $0.59 \pm 0.030$ | $0.57 \pm 0.016$ | $0.59 \pm 0.027$ | $0.67 \pm 0.027$ | $0.58 \pm 0.021$ | $0.68 \pm 0.027$ | $0.55 \pm 0.030$ | $0.59 \pm 0.029$ |
| XGBoost (opt) | $0.64 \pm 0.036$ | $0.63 \pm 0.025$ | $0.63 \pm 0.035$ | $0.72 \pm 0.028$ | $0.60 \pm 0.023$ | $0.72 \pm 0.030$ | $0.57 \pm 0.019$ | $0.64 \pm 0.034$ |

#### 3.2.2 Racial information in oxygen saturation values measured using pulse oximetry

We also investigated the presence of racial information in oxygen saturation values stemming from measurement technologies by comparing performance of race prediction using oxygen saturation values obtained from pulse oximetry (non-invasive) with those obtained from an arterial line (invasive).

Our findings show that pulse oximetry is a significant source of racial information with an AUC of 0.72 (± 0.028). This is in contrast with the results from invasively measured oxygen saturation (arterial line), where these values predict patients’ self-reported race with AUC of 0.60 (± 0.023), as shown in Table 3. Our results support the previous findings that pulse oximetry is affected by the colour of the skin ^16^.

### 3.3 Racial information in care delivery practices reflected in vital signs

Additionally, we focused on care delivery practices as reflected in the frequency of measurements of vital signs. Therefore, we used the frequency of measurement of vital signs, rather than the actual values. We find that care delivery practices do not significantly influence prediction of patients’ race with an AUC of 0.57 (± 0.019), as shown in Table 3. This may also be because vital signs are measured far more routinely than other variables, and consequently if bias indeed exists it would be difficult for the algorithms to ascertain.

### 3.4 Other sources of racial information reflected in patients’ vital signs

Finally, we investigated whether self-reported racial information can be learned even when controlling for measurement technologies and care delivery practices. For this analysis we used the values of invasively measured vital signs only. This is because invasively measured vital signs are less prone to be influenced by measurement technology. Even when controlling for these factors, we showed that self-reported race can be learned from vital signs with an AUC of 0.64 (± 0.034), as shown in Table 3.

Without further investigation, it is difficult to pin-point potential sources of self-reported race. One hypothesis could be calibration differences in APACHE IV scores as shown in ^17^, or differences in disease severity not being reflected in the APACHE IV scores ^9^. However, upstream factors, such as patient selection criteria for an arterial line or even availability of patients might have also contributed to self-reported race being embedded in vital signs ^18^.

## 4 Discussion

We have shown that machine learning algorithms can learn self-reported racial information from vital signs alone. This is unexpected as racial information was not thought to be present within values of vital signs. Furthermore, the ability of machine learning algorithms to learn racial information from vital signs generalised to diverse patient populations and held even when controlling for known sources of racial bias, such as pulse oximetry readings affected by skin colour, care delivery practices, which were not found to contribute to racial information, and presence of co-morbidities.

Learning self-reported racial information creates a significant concern for development of algorithms that support clinical decision-making. If sensitive attributes such as race are learned while training algorithms to predict or optimise an outcome, it is possible that the algorithms will use these features to inform decision-making. Even more concerning is when these sensitive attributes are learned from essential clinical information, heavily relied upon to inform not only course of treatment, but also operational workflows and policy making.

There is overwhelming evidence that race factors into clinical decision-making^19^. Hispanic patients seen by non-Hispanic providers received breast and colorectal cancer screening at higher rates than Hispanic patients seen by Hispanic providers^20^. Greenwood and colleagues reported a 58% reduction in mortality of Black newborns when they are under the care of Black physicians compared to when they are under the care of White physicians^21^. Despite reporting greater pain and pain-related disability, minority patients are more likely to receive inadequate pain treatment compared with white patients^22–24^. When a sensitive attribute such as race is learned even when deliberately removed from a dataset, then it is not far-fetched to think that algorithms will also pick up provider bias and subjectivity in decision-making. After all, algorithms are trained on data that document clinical intuition and judgement, encoded as decisions that contribute to outcome disparities across race, sex, and other demographic factors.

While definite prediction of self-reported race cannot be obtained, the success of the model in correctly classifying two out of three patients is not accidental. Our machine learning models are using only the information from patients’ vital signs, suggesting that statistical features from routinely collected information in the first 24 hours of admission contain embedded information along racial dimensions. Pulse oximeter readings, considered an important unbiased measure of hypoxemia, were shown to be influenced by skin colour, which came to light during the COVID crisis ^25–28^. Upon further investigation, it was revealed that oxygen saturation levels had greater variability in patients who identified as Black, followed by Hispanic, Asian American, and least in White patients. While our saliency analysis showed pulse oximetry as an important variable, its correlation with self-reported race does not fully explain our findings.

In addition to the potential risks, our study also highlights challenges in mitigation measures. A common approach, although criticised^29^, is to selectively remove variables that encode sensitive attributes, such as that AI models do not learn from them and consequently do not become part of the decision process. Ubiquitous use of vital signs in clinical decision-making renders this approach impossible, not least because the origin of racial information appears to be difficult to isolate. Perhaps the time has come to apply a counter approach, by using sensitive attributes such as race to facilitate audit for possible algorithmic bias and adapt established policies on how to ethically collect, use, and report data on race and ethnicity ^30^.

### 4.1 Limitations

While our study includes a large and diverse patient population (335 ICUs in 208 US hospitals), allowing investigation of bias for several racial groups based on an open and well-studied dataset (eICU-CRD), some limitations are present. Use of self-reported race presents a challenge, as studies have shown that genetic variability is higher within the races than between the races ^31^, rendering race a more social construct rather than a biological one. Following on, self-reported race in this study included rigid categories that did not account for patients of mixed ancestry as well as limited availability of data from other racial identity categories. We included Black, Hispanic, and White patients only. Other racial identities (namely Asian American and Native American patients) had insufficient data to properly analyse. Furthermore, there is a discussion in recent years of whether studies are revealing race or results of racism. While this study focuses on the categories of race, the potential explanations given for the findings around race are focused on the impact of racism in society and consequently medicine. Finally, our study focused on US-based patient population, therefore further investigation would be required to determine whether these results are generalisable to centres outside of US-based ICUs.

## 5 Conclusion

As machine learning weaves itself into the fabric of healthcare, there is an increasing attention on the effect of algorithms on underrepresented, marginalised, or disadvantaged populations. For example, algorithms used to identify patients with complex health needs were found to perpetuate racial disparities ^32^, leading to a call for greater algorithmic transparency by the US Senate. Our work, while in the same vein, goes beyond this call by additionally drawing attention to unexpected sources of bias and the potential harm given their ubiquitous use in clinical decision-making.

## Data Availability

All the data for this study is available at https://eicu-crd.mit.edu/gettingstarted/access/

https://eicu-crd.mit.edu/

## 6 Declarations

**Data Sharing**: All data used in this study is publicly available from PhysioNet (https://www.physionet.org/content/eicu-crd/2.0/)

**Author Contributions**: BV, HG, DD and VO had full access to all of the data in the study and take responsibility for the integrity of the data and the accuracy of the data analysis.

**Concept and design**: BV, HG, DD, LAC, VO

**Acquisition, analysis, or interpretation of data**: BV, HG, DD, LAC, VO

**Drafting of the manuscript**: BV, HG, DD, LAC, VO

**Critical revision of the manuscript for important intellectual content**: BV, HG, DD, MK, IDM, JWG, NM, LAC, VO

**Statistical analysis**: BV, HG, DD, VO

**Obtained funding**: HG, LAC, VO

**Conflict of Interest Disclosures**: None

**Funding/Support**: This research was partially supported by the WideHealth project - EU Horizon 2020, under grant agreement No 952279.

**Role of the Funder/Sponsor**: The funder had no role in the design and conduct of the study; collection, management, analysis, and interpretation of the data; preparation, review, or approval of the manuscript; and decision to submit the manuscript for publication.

## Appendix 1 Baseline prediction of self-reported race

**Table 2.** Baseline race prediction performance in the general cohort for Logistic Regression (LR), XGBoost, and XGBoost with optimised (opt) hyperparameters. Results are shown using Area Under the receiver operating characteristic Curve (AUC) along with standard deviation (SD).

|  | Black and White patients | Hispanic and White patients | Hispanic and Black patients |
| --- | --- | --- | --- |
| LR | 0.61 ± 0.029 | 0.61 ± 0.048 | 0.70 ± 0.056 |
| XGBoost | 0.73 ± 0.034 | 0.66 ± 0.079 | 0.78 ± 0.083 |
| XGBoost (opt) | 0.74 ± 0.022 | 0.63 ± 0.098 | 0.75 ± 0.092 |

## Appendix 2 cohort selection diagram

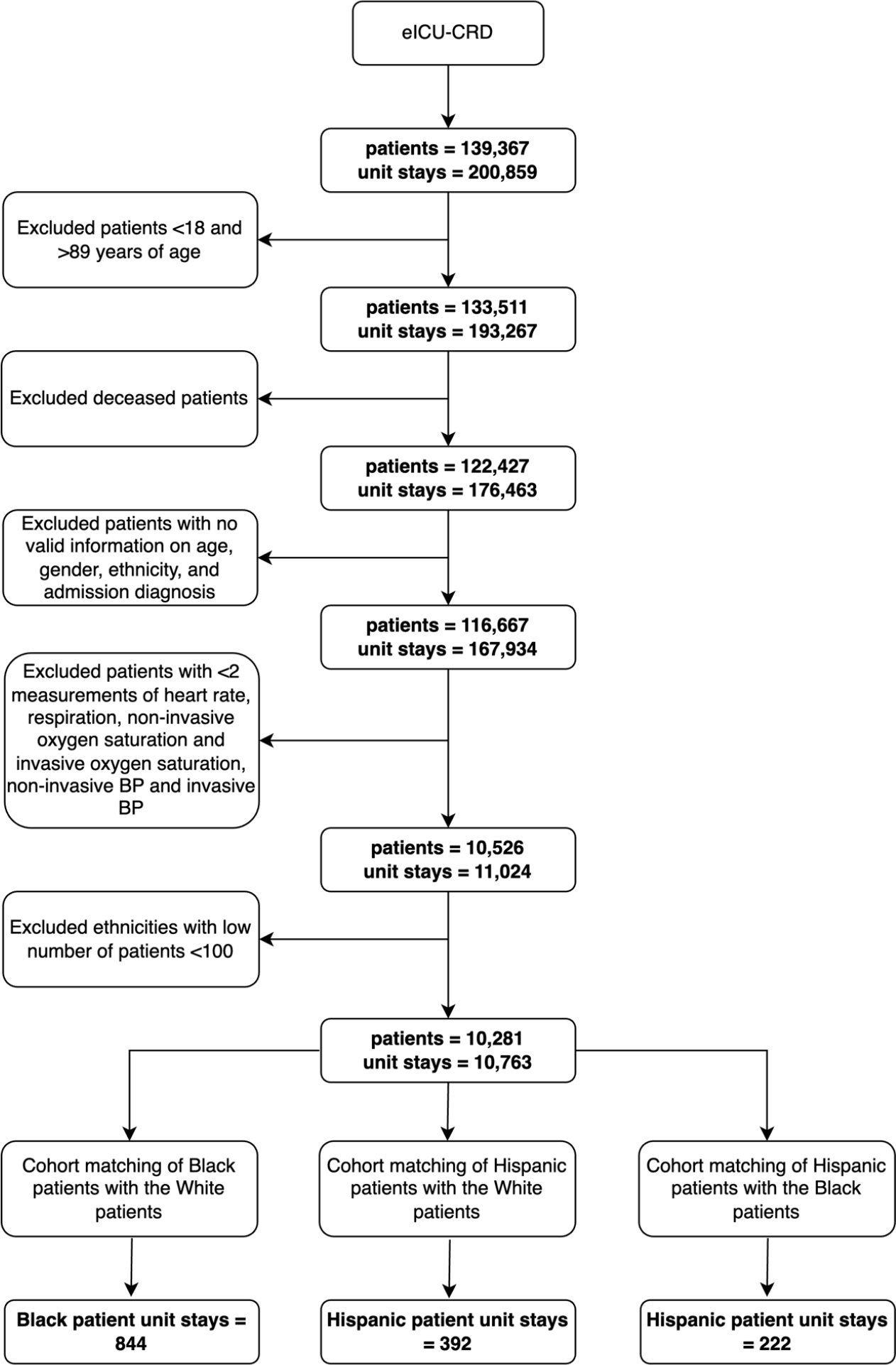

